# Cash versus Lotteries: COVID-19 Vaccine Incentives Experiment*

**DOI:** 10.1101/2021.07.26.21250865

**Authors:** Raymond M. Duch, Adrian Barnett, Maciej Filipek, Laurence Roope, Mara Violato, Philip Clarke

## Abstract

Governments are considering financial incentives to increase vaccine uptake to end the COVID-19 pandemic. Incentives being offered include cash-equivalents such as vouchers or being entered into lotteries. Our experiment involved random assignment of 1,628 unvaccinated participants in the United States to one of three 45 second informational videos promoting vaccination with messages about: (a) health benefits of COVID-19 vaccines (control); (b) being entered into lotteries; or (c) receiving cash equivalent vouchers. After seeing the control health information video, 16% of individuals wanted information on where to get vaccinated. This compared with 14% of those assigned to the lottery video (odds ratio of 0.82 relative to control: 95% credible interval 0.57-1.17) and 22% of those assigned to the cash voucher video (odds ratio of 1.53 relative to control: 95% credible interval 1.11-2.11). These results support greater use of cash vouchers to promote COVID-19 vaccine uptake and do not support the use of lottery incentives.

---

In an effort to control the propagation of the COVID-19 virus, governments world-wide are rolling out aggressive vaccination campaigns. The goal is to vaccinate a sufficient portion of their populations in order to ensure “herd immunity” - or at least to allow life and economic activity to return to normal without a severe burden to population health and health care systems. Policy makers, guided by the scientific community, have anticipated that herd immunity will require COVID-19 vaccination rates of up to 80 percent of the population [4, 1]. While there is now uncertainty over not only the precise level of this immunity threshold, but also whether it is even achievable, much higher vaccination rates than the current 56% of the US population (or two thirds of adults) will be needed to allow life to return to normal without ongoing crippling health and economic effects of of the virus. It is likely that countries, in spite of having sufficient supplies of vaccine, will fall short of these goals. In the U.S., where at least 40 percent of the adult population have still not received at least one vaccination, COVID-19 vaccination rates slowed, from a peak of 3.38 million shots on April 13, 2021, to fewer than 2 million doses per day in May [45]. Contributing to this challenge is the fact that large percentages of many national populations continue to express reservations regarding COVID-19 vaccination [22]. Governments are considering different policies to encourage vaccination of these segments of the population that are either hard to reach or difficult to convince.

Messaging plays a critical role in these efforts to promote COVID-19 vaccination. In the early stages of the COVID-19 vaccine campaigns government messaging has relied, for example, on appeals to social responsibility, endorsements by prominent citizens, reassurances about vaccine brands, data on vaccine efficacy, and allaying the public’s health concerns. There is considerable research quantifying the impact of these different messaging themes on vaccine uptake [3, 2].

However, appeals to medical self-interest or to a vaccine “social contract” [25, 47, 38] have not resonated with all non-vaccinated segments of the population. In a number of countries vaccine campaigns have begun to implement various types of monetary incentives that have been particularly targeted at the hard to convince segments of the non-vaccinated population. Messaging campaigns inform the public that they can earn, or have a chance to earn, cash or cash-equivalents if they get vaccinated. These incentives are typically either cash-equivalents such as online retail vouchers or being entered into a lottery for a prize in the form of either cash or a high-valued item such as a car [42]. Such an approach builds on a wide range of experimental evidence on the use of financial incentives to promote vaccination and other preventive behaviours, with a recent review concluding that “[F]inancial incentives may be a useful addition to the behavioural change toolkit” [14]. Evidence from recent surveys indicates that financial incentives may play a role in increasing rates of COVID-19 vaccination. Financial incentives, rather than information and appeals to the common good (or even personal advantage), convinced experimental subjects to subscribe to a COVID-19 contact tracing app [30]. A Kaiser Family Foundation study found that about one in four would be more likely to get vaccinated if they were entered into a lottery with a chance to win one million dollars [19]. Initial evidence from a German conjoint experiment suggests that a hypothetical financial incentive of 50 Euros, as part of a mass vaccination scenario, could increase vaccine uptake among the hesitant [20]. Similarly, a third of the unvaccinated respondents in a U.C.L.A. COVID-19 Health and Politics Project survey experiment indicated cash payments would make them more likely to get vaccinated [44].

Many U.S. state governments have adopted a variety of cash and lottery incentives in order to encourage citizens to get a COVID-19 vaccine. The National Governors Association has published a description of the various COVID-19 vaccines available in the different U.S. states [32]. While these schemes have garnered considerable enthusiasm amongst policy makers, a number of reservations have been raised regarding their morality, efficacy and possible negative consequences [45, 23, 21]. A recent study of vaccination rates before and after the implementation of lottery incentives in Ohio did not find significant increases in adult COVID-19 vaccination rates [46]. This has led some [13] to argue for abandoning lotteries in favour of other approaches including cash vouchers, which have been shown to be more effective than lottery incentives for other preventative behaviours such as chlamydia screening [33]. However, prominent scholars, such as Richard Thaler, have argued that lotteries are an effective way to increase vaccination rates [39, 34]. There is an evidence gap with respect to incentives. While we have abundant experimental clinical evidence on the effectiveness of COVID-19 vaccines, surprisingly, we have limited experimental evidence informing policies designed to encourage COVID-19 vaccine uptake.

This study conducts a randomized experiment to inform the policy debate regarding the efficacy of cash and lotteries as incentives for individuals to get a COVID-19 vaccination. Vaccine incentive policies have varied greatly across U.S. states, which facilitated the design of both the informational treatment videos and the outcome measure [24]. We implemented a randomized experiment to determine which of three messaging strategies is most effective for encouraging the non-vaccinated to seek information on how to get vaccinated: a standard CDC-inspired message that identifies the personal and public health benefits of COVID-19 vaccination; a message that highlights the chance of winning a lottery if vaccinated; and finally a message that indicates that first-time vaccinated would receive a cash voucher. Our approach was based on the assumption that a basic public health statement stressing the importance, efficacy and safety of COVID-19 vaccines would occur in any real-world information provision. We therefore tested the effects of additional pieces of information that highlighted the availability of cash and lottery incentives for being vaccinated.

The online sample of just over 1,500 non-vaccinated subjects were randomly assigned (1:1:1) to the informational treatment and control videos. After viewing the assigned video, participants were given the opportunity to consult additional information on being vaccinated in their state. We treat this digital expression of interest as our outcome variable. We had two primary preregistered hypotheses: 1) Participants in Video Treatment 2 (lottery message) would be more likely to click through to the vaccination information web page than participants assigned to the Treatment 1 control group (standard CDC-inspired health message); 2) Participants in Video Treatment 3 (cash voucher message) would be more likely to click through to the vaccination information web page than participants assigned to the Treatment 1 control group (standard CDC-inspired Health message). Our secondary hypothesis was that there would be no difference in click through rates between Treatment 2 and Treatment 3.^1^

## Results

Figure 1 describes the details of the sampling and treatment assignment. A total of 3,416 online participants were recruited to participate in the CANDOUR Incentive study (3,551 from the Cloud Research Prime Panel; 297 recruited via Facebook; and 364 recruited from Lucid Fulcrum Exchange). For the 1,798 participants who indicated that they had received at least one COVID-19 vaccine the Incentive survey terminated. The 1,618 participants who indicated they had not had a COVID-19 vaccination were then invited to complete the online survey.^2^ the Online participants were paid 2 USD for the experiment, which lasted less than 5 minutes. At the outset of the survey participants were asked five brief demographic questions (the full survey is available in the Online Supplemental Materials). Participants were then randomly assigned to one of the three video messages with the pre-registered objective of approximately 500 participants per video. We implemented sequential covariate-adaptive randomization. For each incoming respondent we calculate the Mahalanobis distance for four key covariates to ensure the random assignment resulted in balance across the three treatment videos [29]: race, gender, age and education. As each new participant entered the online experiment we adjusted the treatment assignment probabilities to ensure balance.^3^.

**Figure 1:**
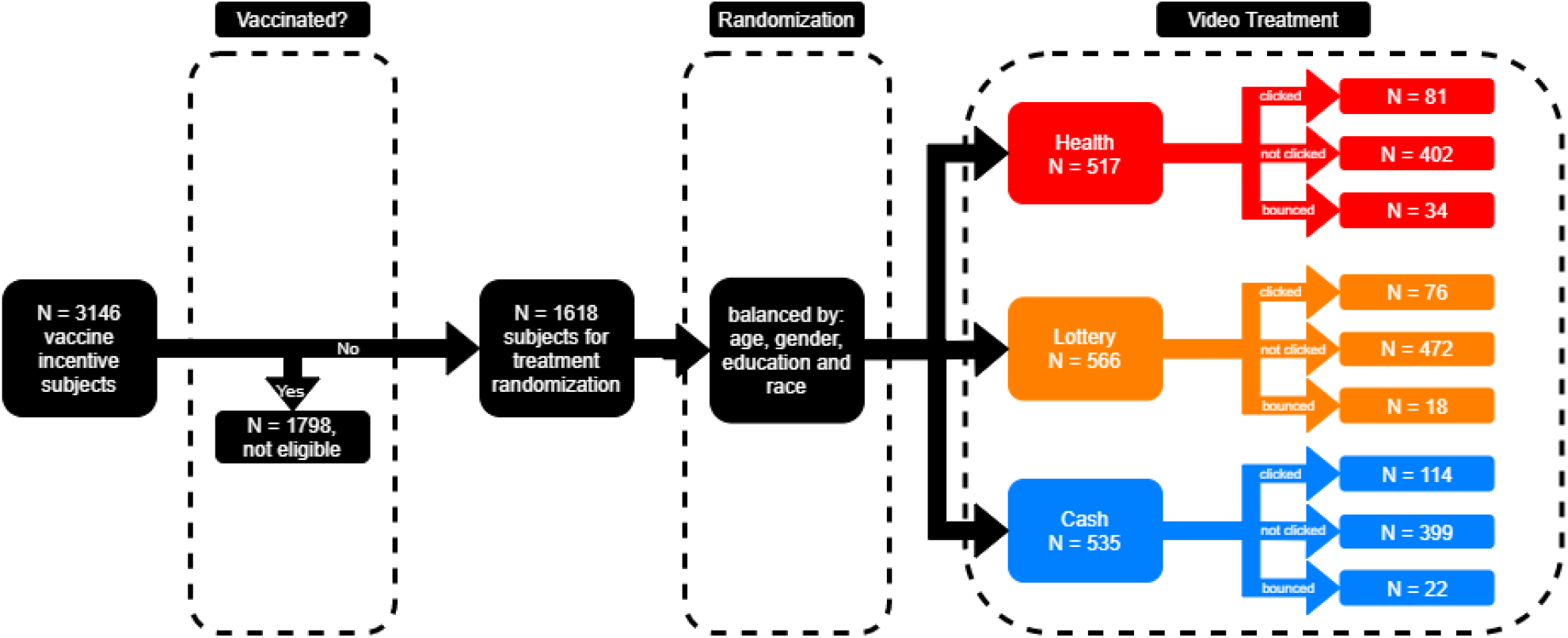
Experiment Flow Diagram

The three information treatments were delivered in a short 45 second video. Treatment 1 (Control): Standard CDC-inspired COVID-19 vaccine promotional and information video. Treatment 2: Lottery treatment – the first 30 seconds are identical to the control video – the last 15 seconds inform viewers that in some states you can be entered into a lottery and win over 1 million dollars if you get vaccinated. Treatment 3: Cash voucher treatment – the first 30 seconds are identical to the control video – the last 15 seconds inform viewers that in some states you can earn money or a money equivalent of up to $ 100 that can be spent online or in stores.^4^ Our treatment design simulates real world decision making situations in which videos play an increasingly important role in delivering information related to public policy [18, 6, 15, 31].

We pre-specified a single primary outcome measure – the digital expression of further interest in vaccination information. After subjects viewed the assigned treatment video, they were provided with a choice between 1) ending the survey or 2) obtaining further information about how they can get vaccinated in their state. The outcome measure is their response to this simple choice. If they clicked on the link for further information, they were directed to this webpage, that we prepared, and which points to vaccination information sources in each state: http://www.didyougetcovidvaccine.com.^5^ This approach builds on online experimental designs that treat digital traces as outcome measures [8, 35, 40, 16]. Their principal advantage is that individuals are not asked to give opinions which can exaggerate experimenter demand effects [36, 31].

Table 1 summarizes the distribution of the covariates for each of the three treatment groups. In addition to age, gender, race and education, we asked state of residence. The percentage of each state’s vote that was won by President Donald Trump in the 2020 presidential race is employed to generate a state Trump vote variable [26]. The state Trump variable scales the state Trump vote share by subtracting the mean percentage and dividing this by 10 percent (State % Trump vote (+10%)). We employ a scaled age variable in the analysis – subtracting 40 from the observed age and dividing the result by 10 (Age (+10 years)). Generally, relative to the U.S. population, the online convenience sample is younger, more white and disproportionately female. The distribution of these covariates within each of the three treatment groups suggests that the non-vaccinated subjects were randomly allocated across the three treatments.

**Table 1:** Summary Statistics for U.S. Vaccine Incentives Treatment Assignment

|  | CDC Health | Cash Voucher | Lottery |
| --- | --- | --- | --- |
| **Education** |  |  |  |
| High | 91 (18%) | 73 (14%) | 93 (17%) |
| Medium | 224 (43%) | 238 (45%) | 230 (41%) |
| Low | 198 (38%) | 219 (41%) | 232 (41%) |
| Missing | 3 (1%) | 1 (0%) | 7 (1%) |
| **Race** |  |  |  |
| Black | 98 (19%) | 84 (16%) | 101 (18%) |
| Other | 33 (6%) | 38 (7%) | 54 (10%) |
| White | 385 (75%) | 409 (77%) | 407 (72%) |
| **Gender** |  |  |  |
| Female | 359 (70%) | 345 (65%) | 363 (65%) |
| Male | 157 (30%) | 185 (35%) | 198 (35%) |
| Other | 0 (0%) | 1 (0%) | 1 (0%) |
| **Pool** |  |  |  |
| CloudResearch | 457 (89%) | 472 (89%) | 492 (88%) |
| Facebook | 17 (3%) | 16 (3%) | 26 (5%) |
| Lucid | 42 (8%) | 43 (8%) | 44 (8%) |
| **Continuous Measures**<br>**(Median and Inter-quartile range)** |  |  |  |
| Age | 36 (29 to 49) | 38 (29 to 46) | 36 (29 to 46) |
| State % Trump vote | 51 (45 to 57) | 51 (44 to 55) | 51 (44 to 53) |
| **Outcome: Click through rates** |  |  |  |
| Frequency | 81 (16%) | 114 (22%) | 76 (14%) |
| Odds Ratio (CI) |  | 1.47 (1.07 to 2.02) | 0.84 (0.60 to 1.18) |
| TOTAL N | 516 | 531 | 562 |

**Table 2:** Multinomial Logit Regression of Treatment Assignments

|  | **CDC Health** | **Lottery** |
| --- | --- | --- |
| Male | -0.23<br>(-0.49 to 0.03) | 0.00<br>(-0.25 to 0.25) |
| Age (+10 years) | 0.03<br>(-0.06 to 0.12) | -0.03<br>(-0.12 to 0.06) |
| Race (Other) | -0.35<br>(-0.90 to 0.21) | 0.17<br>(-0.34 to 0.68) |
| Race (White) | -0.24<br>(-0.58 to 0.09) | -0.15<br>(-0.48 to 0.18) |
| Education (Low) | -0.35<br>(-0.72 to 0.02) | -0.18<br>(-0.55 to 0.18) |
| Education (Medium) | -0.32<br>(-0.68 to 0.04) | -0.27<br>(-0.63 to 0.09) |
| State % Trump vote (High) | 0.14<br>(-0.10 to 0.39) | 0.02<br>(-0.22 to 0.26) |
| Note: Multinomial coefficients reported with 95% confidence intervals |  |  |

### Cash incentives, not lotteries, appeal to the non-vaccinated

Table 1 presents the click through rates for participants assigned to the three treatments. Of the 516 participants assigned to the control, i.e. the CDC-inspired health video, 81 (16%) subsequently expressed interest in more vaccine information. A slightly lower 14% of those assigned to the lottery video subsequently clicked on the link for further information. The treatment odds ratio for this lottery video is 0.82 (95% credible interval of 0.57 to 1.17, p-value = 0.27). Hence, there is no compelling evidence that lottery incentives perform any better than a standard health message. A much higher 22% of participants assigned to the cash voucher video subsequently clicked through to the further information page. With an odds ratio of 1.53 (95% credible interval of 1.11 to 2.11, p-value = 0.009), the odds of these participants expressing interest in information is about 1.5 times greater than the odds of the control subjects.

We estimate a Bayesian logistic regression model with a dichotomous dependent variable indicating whether the respondent clicked through for more vaccine information. Table 3 in Methods presents the results for a basic specification with the Cash and Lottery treatment variables (Model 1); a Model 2 that includes the full set of covariates; and a Model 3 that adds random effects for States and participant pools.

**Table 3:**
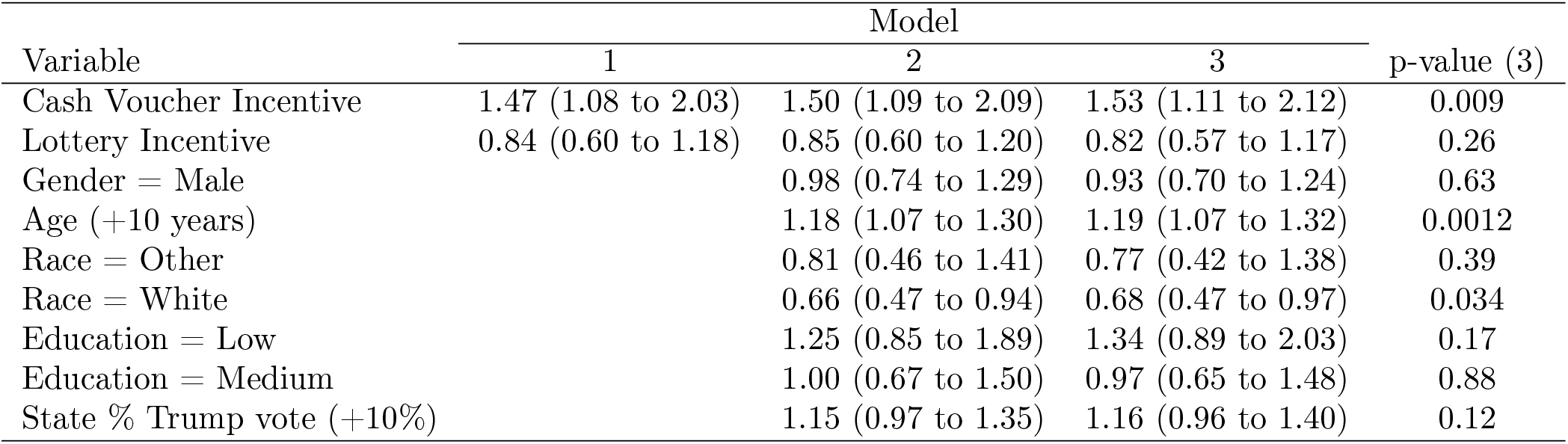
Odds ratios and 95% credible intervals from three multiple variable Bayesian logistic regression models of the odds of click-through

| Variable | Model |  |  | p-value (3) |
| --- | --- | --- | --- | --- |
|  | 1 | 2 | 3 |  |
| Cash Voucher Incentive | 1.47 (1.08 to 2.03) | 1.50 (1.09 to 2.09) | 1.53 (1.11 to 2.12) | 0.009 |
| Lottery Incentive | 0.84 (0.60 to 1.18) | 0.85 (0.60 to 1.20) | 0.82 (0.57 to 1.17) | 0.26 |
| Gender = Male |  | 0.98 (0.74 to 1.29) | 0.93 (0.70 to 1.24) | 0.63 |
| Age (+10 years) |  | 1.18 (1.07 to 1.30) | 1.19 (1.07 to 1.32) | 0.0012 |
| Race = Other |  | 0.81 (0.46 to 1.41) | 0.77 (0.42 to 1.38) | 0.39 |
| Race = White |  | 0.66 (0.47 to 0.94) | 0.68 (0.47 to 0.97) | 0.034 |
| Education = Low |  | 1.25 (0.85 to 1.89) | 1.34 (0.89 to 2.03) | 0.17 |
| Education = Medium |  | 1.00 (0.67 to 1.50) | 0.97 (0.65 to 1.48) | 0.88 |
| State % Trump vote (+10%) |  | 1.15 (0.97 to 1.35) | 1.16 (0.96 to 1.40) | 0.12 |

Figure 2 plots the odds ratios and their credible intervals for the fully specified Model 3. The estimated odds ratios for Cash and Lottery in Model 1 are, respectively, 1.47 (95% credible interval of 1.08 to 2.03) and 0.84 (95% credible interval of 0.60 to 1.18) and they vary little across all three model specifications. Again, this confirms that the odds of participants in the Cash Video Treatment of expressing interest in information are about 1.5 greater than the odds of the control participants who see a standard CDC-inspired Health Video. Those in the Lottery Video Treatment have an estimated odds ratio with credible confidence intervals that encompass 1 – providing little evidence that the lottery had any benefit. Note also that the credible intervals on the Cash Voucher and Lottery odds ratios just barely intersect – reasonable evidence that Cash rather than Lotteries appeals to the non-vaccinated.

**Figure 2:**
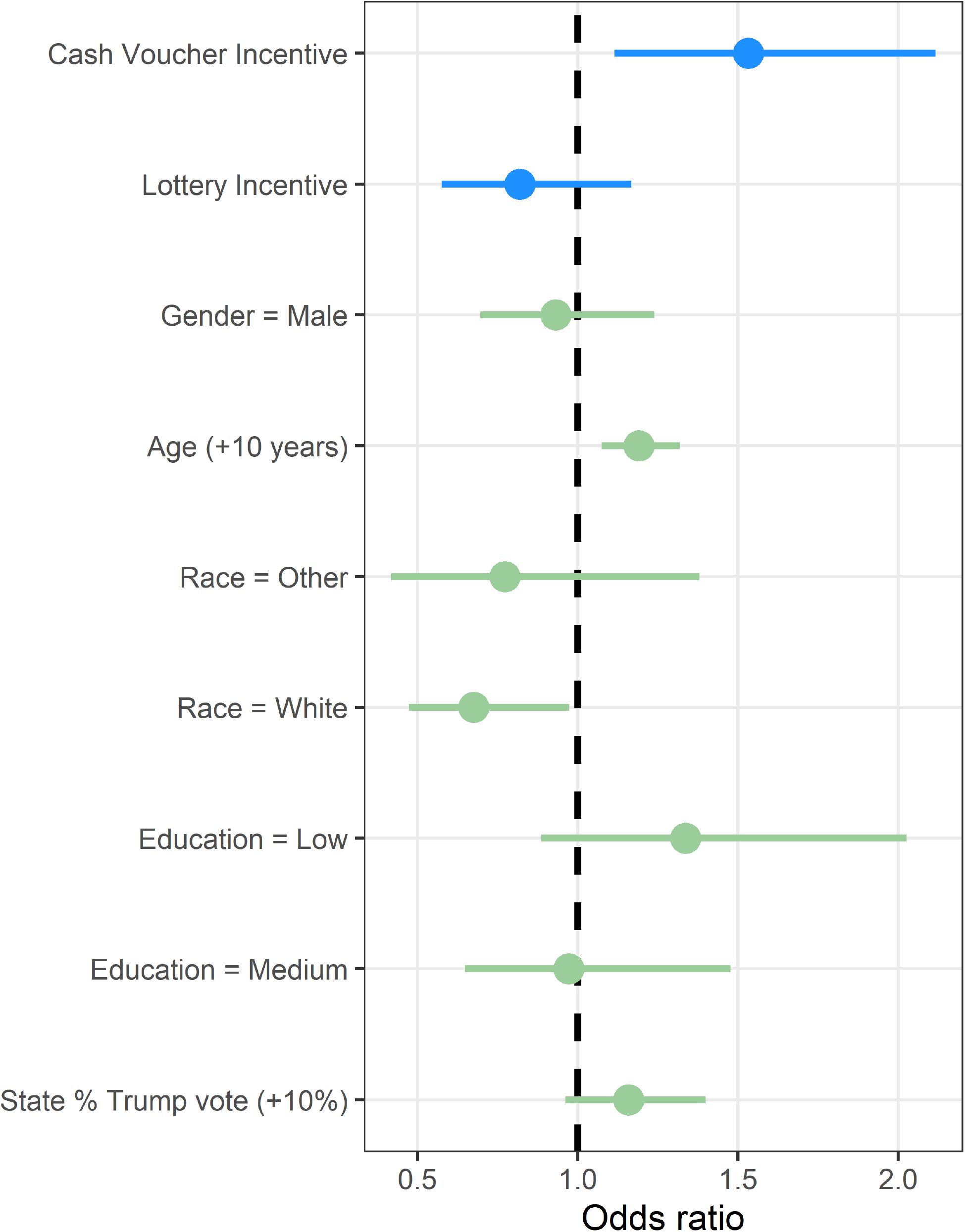
Bayesian Logistic Regression Model of Clicks to Vaccine Information. Odds ratios and 95% credible intervals.

Estimated odds ratios for the covariate adjustments, from Model 3 in Table 3 in Methods, are also presented in Figure 2. Two of the covariate adjustments are statistically significant. Whites have an odds ratio of 0.68 (95% credible interval of 0.47 to 0.97) indicating they were less likely than Blacks to click through for more information after the video treatment. Age has an odds ratio of 1.19 per 10 year increase (95% credible interval of 1.07 to 1.32) indicating older participants were more likely to click through for more vaccine information.

### Heterogeneity

We specified heterogeneous treatment effects for three covariates: gender, race and education. The specifications are presented in Equations 1 to 4 (Methods). The interaction terms in these models indicate whether exposure to one of the two treatments (Cash or Lottery) induces an increase in vaccination information interest, relative to the control in that same subgroup. Hence, in the case of education, we present the relative effect of changing treatment within the same education level: High Education and Cash vs High Education and Control; Medium Education and Cash vs Medium Education and Control; Low Education and Cash versus Low Education and Control. In Tables 4 to 6 (Methods), we present the estimated odds ratios with 95% credible intervals and Bayesian p-values. In Figure 3, we present these estimated odds ratios along with their credible intervals for Education, Gender and Race.

**Table 4:** Odds ratios and 95% credible intervals for the multiple logistic regression model examining an interaction between gender and treatment

| Variable | OR | CI | p-value |
| --- | --- | --- | --- |
| Cash Voucher Incentive | 1.83 | 1.23 to 2.75 | 0.004 |
| Lottery Incentive | 0.97 | 0.63 to 1.49 | 0.899 |
| Gender = Male | 1.33 | 0.79 to 2.22 | 0.279 |
| Male × cash voucher | 0.59 | 0.29 to 1.19 | 0.131 |
| Male × lottery | 0.61 | 0.28 to 1.28 | 0.186 |
| Age (+10 years) | 1.19 | 1.08 to 1.32 | <0.001 |
| Race = Other | 0.75 | 0.41 to 1.34 | 0.351 |
| Race = White | 0.67 | 0.47 to 0.96 | 0.030 |
| Education = Low | 1.34 | 0.90 to 2.03 | 0.159 |
| Education = Medium | 0.98 | 0.65 to 1.48 | 0.902 |
| State % Trump vote (+10%) | 1.16 | 0.96 to 1.41 | 0.114 |

**Table 5:** Odds ratios and 95% credible intervals for the multiple logistic regression model examining an interaction between race and treatment

| Variable | OR | CI | p-value |
| --- | --- | --- | --- |
| Cash Voucher Incentive | 1.60 | 1.10 to 2.31 | 0.014 |
| Lottery Incentive | 0.71 | 0.46 to 1.07 | 0.108 |
| Race = Other | 1.32 | 0.49 to 3.31 | 0.551 |
| Race = Black | 1.22 | 0.63 to 2.28 | 0.547 |
| Other race × cash voucher | 0.72 | 0.19 to 2.60 | 0.610 |
| Other race × lottery | 0.89 | 0.24 to 3.22 | 0.868 |
| Black race × cash voucher | 0.88 | 0.36 to 2.08 | 0.782 |
| Black race × lottery | 2.06 | 0.86 to 4.92 | 0.110 |
| Age (+10 years) | 1.19 | 1.08 to 1.32 | <0.001 |
| Gender = Male | 0.91 | 0.68 to 1.22 | 0.531 |
| Education = Low | 1.32 | 0.87 to 2.01 | 0.195 |
| Education = Medium | 0.97 | 0.64 to 1.46 | 0.856 |
| State % Trump vote (+10%) | 1.16 | 0.96 to 1.41 | 0.115 |

**Table 6:** Odds ratios and 95% credible intervals for the multiple logistic regression model examining an interaction between education and treatment

| Variable | OR | CI | p-value |
| --- | --- | --- | --- |
| Cash Voucher Incentive | 1.47 | 0.89 to 2.47 | 0.130 |
| Lottery Incentive | 0.72 | 0.42 to 1.24 | 0.242 |
| Education = Medium | 0.59 | 0.34 to 1.02 | 0.061 |
| Education = High | 0.84 | 0.42 to 1.68 | 0.633 |
| Medium education × cash voucher | 1.16 | 0.55 to 2.36 | 0.688 |
| Medium education × lottery | 1.67 | 0.77 to 3.60 | 0.192 |
| High education × cash voucher | 1.01 | 0.39 to 2.61 | 0.983 |
| High education × lottery | 0.58 | 0.18 to 1.72 | 0.342 |
| Age (+10 years) | 1.19 | 1.08 to 1.32 | <0.001 |
| Gender = Male | 0.93 | 0.69 to 1.24 | 0.622 |
| Race = Other | 0.76 | 0.41 to 1.35 | 0.364 |
| Race = White | 0.67 | 0.47 to 0.98 | 0.041 |
| State % Trump vote (+10%) | 1.16 | 0.97 to 1.40 | 0.109 |

**Figure 3:**
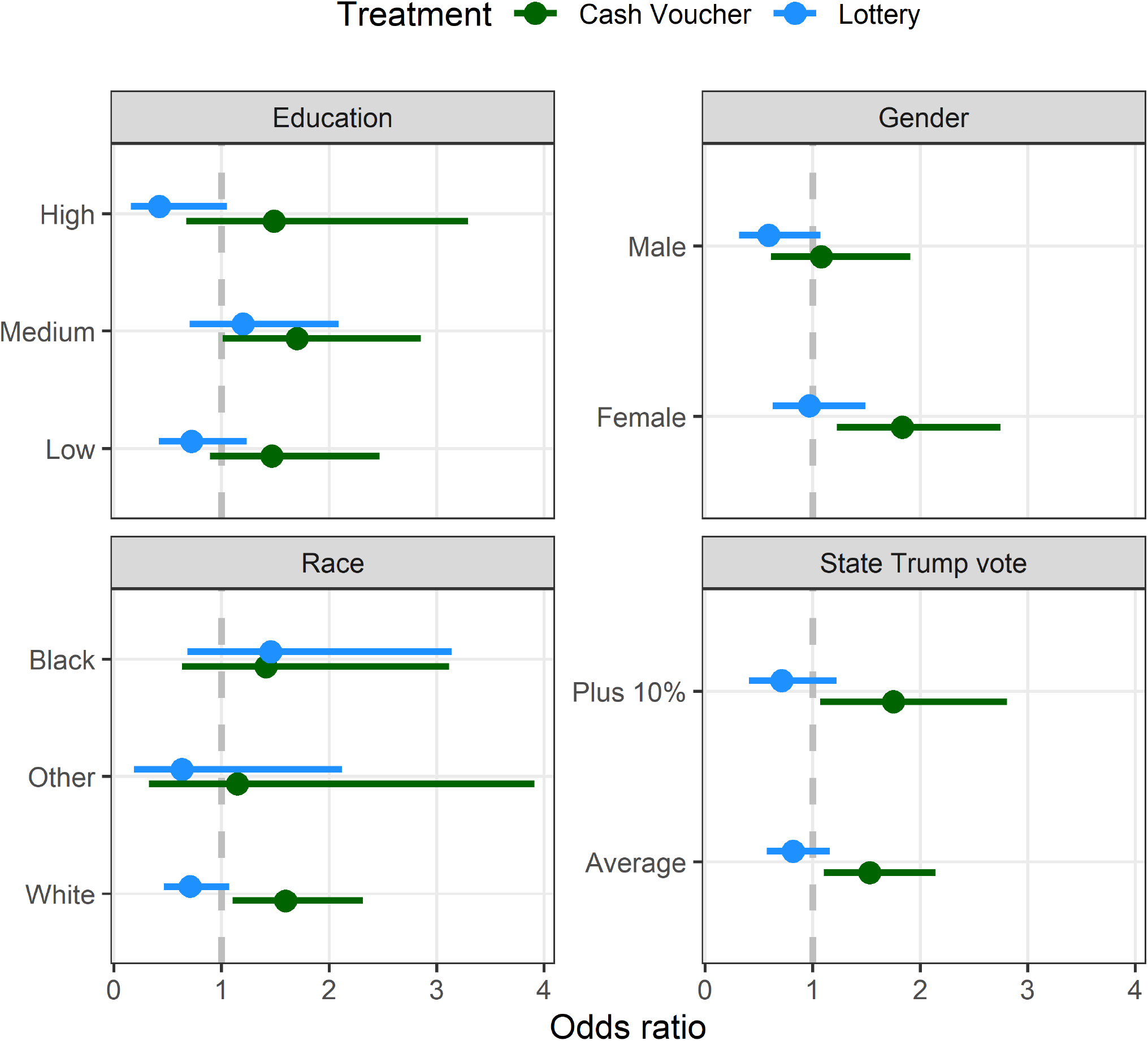
Estimated odds ratios and 95% credible intervals of the treatment effect interacted with with gender, race, education and state Trump vote from the Bayesian Logistic Regression Model

The set of three Education and Cash Voucher odds ratios are similar to the overall Cash odds ratio of 1.5. The Education and Lottery odds ratios are consistent with our overall findings – the Lottery treatment effect for the most part is indistinguishable from the control. There is some evidence in Figure 3 that the Lottery treatment effect for the High Education group is significantly lower than the control treatment for the highly educated. There is a gender interaction. For female participants, the Cash Voucher odds ratio with respect to the control is approximately 1.8 and quite precisely estimated, while for male participants the odds ratio is close one.

The race interactions suggest interesting differences between Whites and Blacks. For Whites, the Cash Voucher odds ratio is approximately 1.5 and precisely estimated (they are the dominant racial group in the sample). The Lottery odds ratio for Whites is less than 1.0 and also precisely estimated. Relative to the control Health video, Whites are clearly more likely to click through for more information when they view the Cash Voucher information treatment and less likely when they view the Lottery treatment. Blacks, on the other hand, appear to respond similarly to both the Cash Voucher and Lottery treatments. The Cash Voucher and Lottery odds ratios, relative to the control, are similar for Blacks – approximately 1.4 in both cases although imprecisely estimated given that Blacks make up a smaller proportion of the sample – approximately 20 percent.

Figure 3 also estimates the odds ratios for Cash versus Control for respondents in a state with an average Trump vote share and Cash versus Control odds ratios for those in a state with a Trump vote share that is 10% above the average. State partisanship does not seem to interact with the treatment effects: participants in states with a relatively high Trump 2020 vote share responded to the Cash and Lottery treatments similarly to those in states with an average Trump 2020 vote share.

## Discussion

Based on an online randomized experiment conducted on unvaccinated adults in the United States, we find that information about cash and lottery incentives have markedly different impacts on COVID-19 vaccine information seeking behaviour. Compared with a brief COVID-19 vaccine informational video message, information on the potential to receive a cash voucher (of up to 100 dollars) for being vaccinated increased the proportion of individuals seeking information (from 16 to 22 percent). Vaccine information seeking behavior in response to a lottery incentive was not significantly different from the non-incentivized COVID-19 health video.

Incentives for vaccinations have attracted increasing interest as many countries have experienced a tailing-off of vaccinations below levels deemed necessary either for herd immunity or suppression of the virus to an extent that would permit life to return to normal. Governments have been trying a variety of incentives including cash or near cash vouchers, vaccine lotteries for cash prizes and for products including cars and even guns [42]. Our U.S. experiment suggests that incentives to be vaccinated can be effective, but that the type of incentive matters – at least in the U.S., cash is effective while lotteries are not.

Achieving greater compliance with the COVID-19 vaccination campaigns is an urgent public health challenge. Experimental research that builds on this simple design can inform policy efforts attempting to advance this goal. Vaccine incentives may, or may not, be an appropriate policy tool in many other countries facing this COVID-19 vaccination challenge. Our experimental design offers a relatively efficient approach to answering this question – random assignment of brief videos describing incentives on offer compared to standard health messaging.

Moreover, this simple design can be extended to provide much richer insights into policies that promote vaccination uptake. An obvious design enhancement would be to vary the size of the incentive payments to determine a dose response relationship. This has been observed for incentives for other preventative health behaviours [5]; but also in survey experiments concerning cash payments and COVID-19 vaccine intentions [44]. Our results also suggest that there maybe some heterogeneity in the treatment effects of both cash and lotteries in different sub-groups of the population. For example, there is a significant difference between cash and lotteries among whites, but not blacks. Conducting further experiments with a sample size large enough to detect differences in sub-groups where vaccine uptake is currently at lower level would allow us to assess the cost-effectiveness and equity impacts of different financial incentives.

Our experimental design focuses very narrowly on cash versus lottery financial incentives for COVID-19 vaccinations. There is evidence that a range of other factors affect vaccine uptake, including the convenience of access to vaccination and the personal freedoms associated with proof of vaccination [20]. Some governments, in fact, are imposing increasingly negative incentives (such as the recent announcement of the French government to mandate proof of vaccination for visiting restaurants and cafes [10]). A more informative, although challenging, experimental design would allow us to identify the causal impact of a full range of policy incentives, both financial and non-financial, on COVID-19 vaccination decisions.

A potential limitation of our study – as with most vaccine uptake studies – is that we do not observe the participants’ vaccine decisions. Our outcome measure is their decision to seek out additional vaccination information. An extension of our design that linked information treatments to actual vaccination decisions would be a more powerful design.

The experimental results reported in this article provide some guidance for incentive policies. First, we provide evidence that COVID-19 vaccination messaging that highlights financial incentives will likely have a bigger motivational impact on the non-vaccinated than is the case for standard COVID-19 messaging that focuses solely on the health benefits. Second, the non-vaccinated are significantly more likely to be motivated by messaging that highlights cash voucher incentives than they are by lottery incentives. In fact, lottery messaging is no more effective than standard CDC health messaging.

## Methods

### Ethics

The survey was conducted according to the University of Oxford’s policy for human participants research and approved by the University of Oxford Medical Sciences Interdivisional Research Ethics Committee (MS IDREC) (Approval ID: R72328/RE001). Informed consent was obtained from each participant at the beginning of the survey. The full text of the informed consent form is included in the attached survey.

### Sample

The sample size was determined with formal power calculations that were pre-registered. The pre-registration includes a detailed discussion of the basis for these calculations. There are a limited number of experimental studies comparing similar incentive treatment effects. A recent study by Haisley et al. [17] compared lotteries with cash incentives for completion of health risk assessments and estimated a 0.20 treatment effect between lottery and control. Kluver et al. [20] conducted a conjoint experiment in Germany that explicitly estimated the effect of financial incentives on vaccine intentions for the undecided – their estimated treatment effect was 5 percent. Our recent research, employing similar informational video treatments, estimated a treatment effect of 0.10 between an information treatment and a placebo video control [12]. Our target was to be able to detect a difference in treatment click through rates of 0.10 with a one-sided 1 percent significance level and a power of 80 percent. Our pre-registration indicated this would require a total sample of about 1,000 (roughly 350 respondents in each treatment group). Accordingly, we, conservatively, set our targeted samples sizes at 500 non-vaccinated respondents for each of the CDC Health, Cash Voucher and Lottery video treatments. At the outset of the study, approximately one-third of the U.S. population had not yet had their first COVID-19 vaccine. In order to meet our target of 1,500 non-vaccinated participants we anticipated inviting approximately 4,500 participants.

A total of 4,118 respondents were invited to participate in the online experiment. Primary recruitment was conducted from Cloud Research’s Prime Panels – a total of 3,551. The Cloud Research Prime Panels has a large and diverse pool of online respondents [9]. Additional individuals were also recruited from online Facebook advertisements (297) [48]. The Lucid Fulcrum Exchange provided 364 participants – this is an online participant pool that matches the characteristics and attitudes of other online pools such as MTurk [11]. The survey experiment began on June 28, 2021 and ended July 11, 2021.

### Video Treatments

The three information treatments were delivered in a short video:

- Treatment 1: A 45 second COVID-19 vaccine information video adapted from the U.S. Center for Disease Control (CDC). Video is available here: https://youtu.be/V3DUCC8xnD0
- Treatment 2: Lottery treatment – the first 30 seconds are identical to the CDC-inspired video – the last 15 seconds inform viewers that in some states you can be entered into a lottery and win over 1 million dollars if you get vaccinated. Video is available here: https://youtu.be/QIm_vbpe_go
- Treatment 3: Cash voucher treatment – the first 30 seconds are identical to the CDC-inspired video – the last 15 seconds inform viewers that in some states you can earn money or a money equivalent of up to $ 100 that can be spent on line or in stores. Video is available here: https://youtu.be/gF045EaPj-o

Figure 4 summarizes the overall design elements of the three videos; it includes the specific story board elements employed to produce the three video treatments.

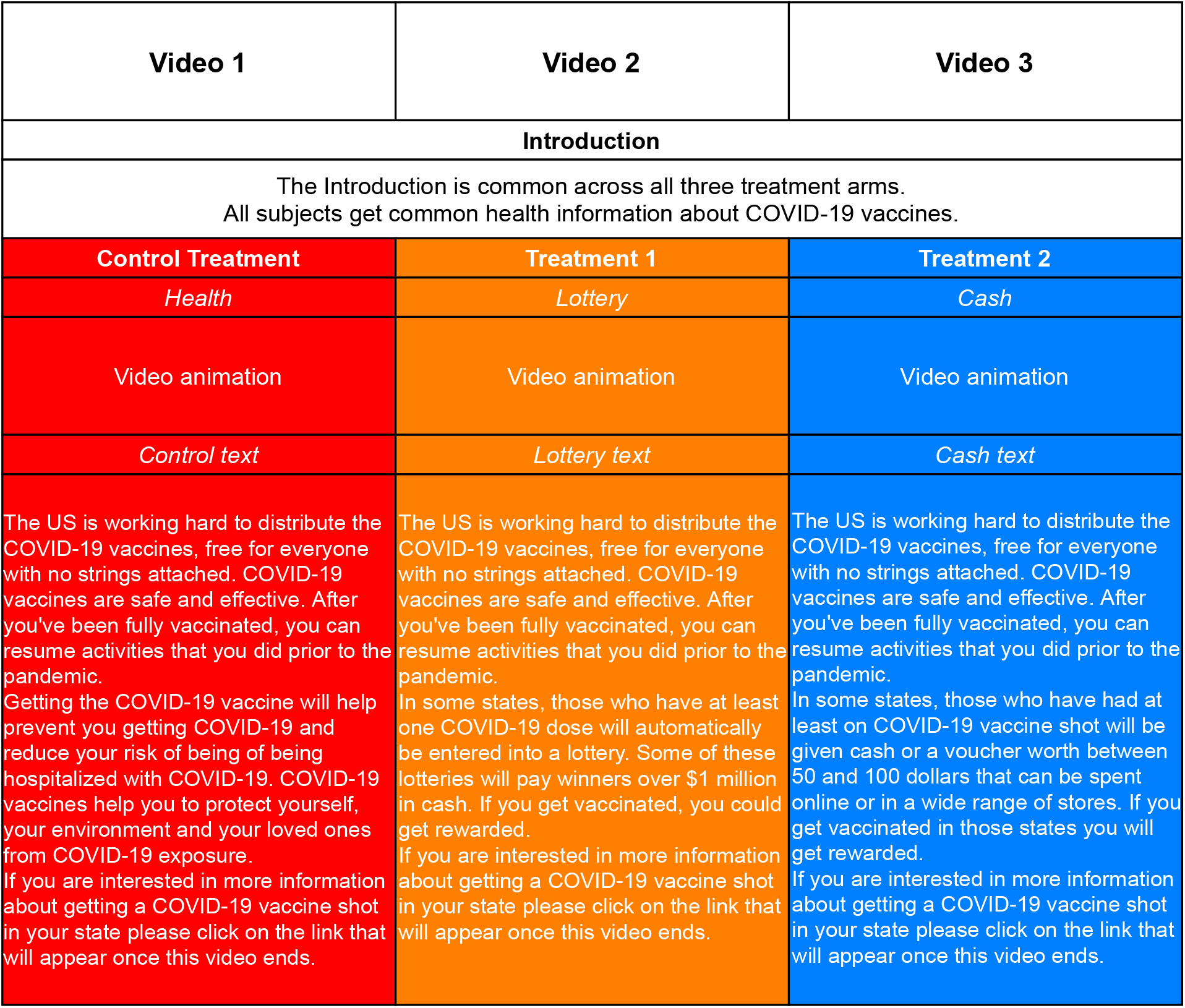

### Treatment Assignment

As indicated in the main text, participants were randomly assigned to one of the three video treatments with the pre-registered objective of approximately 500 participants per treatment. We implemented sequential covariate-adaptive randomization. For each incoming respondent we calculate the Mahalanobis distance for four key covariates to ensure the random assignment resulted in balance across the three treatment videos [29]: race, gender, age and education. As each new participant entered the online experiment, we adjusted the treatment assignment probabilities to ensure balance. Assignment probabilities change based on the covariate profiles of units already in the three treatment groups.

In practice, this procedure seeks to create similar covariate distributions in the treatment groups by biasing the current unit’s treatment assignment in favor of treatment conditions with fewer units with similar profiles. This approach is especially adapted to online experiments such as ours in which we do not know the precise characteristics of respondents who will make up the final sample [43]. At any point in time during the survey period, we only have information on participants who have already taken the survey and the subject that has just arrived.

We implement the sequential covariate adaptive randomization using the seqblock algorithm in the blocktools R package [29, 28, 27, 7]. The adaptive algorithm calculates the Mahalanobis distances and ranks them to bias the randomization toward treatment groups with high scores (the value of Mahalanobis distance). We use the Qualtrics Web Service feature to run this co-variate adaptive randomization remotely using an API.^6^

#### Balance

Table 1 in the text summarized the covariate distributions within each of the three treatment groups. We supplement this by modelling treatment assignment as a function of our five covariates: gender, race, education, age, and State % Trump Vote High (>50%). Table 2 presents the results from a multinomial logit regression with the three treatment assignments (CDC Health, Lottery and Cash Voucher) as the values for the dependent variable. These results simply confirm our earlier observations regarding the covariate balance in treatment assignments we observed in Table 1. As Table 2 indicates, all of the estimated credible intervals encompass zero. This suggests that there is no strong evidence of treatment assignment being correlated with any of our covariates.

### Multiple variable models

We summarize the treatment effects using a Bayesian logistic regression model with click-through as the dependent variable. We used a multiple variable model by including the treatment and potential predictors of click-through (age, gender, race, education and Trump vote). We also included random effects for each state and the three sample pools. We used vague priors for all parameters. In the heterogeneity analyses we included interactions between the treatment effect and the four key variables of: gender, race, education and state % Trump vote.

We present the estimates as odds ratios with 95% credible intervals and Bayesian p-values. The Bayesian p-values estimate the probability that the odds ratio is equal to one. We compared the model fit after adding additional covariates to the logistic regression model using the Deviance Information Criterion (DIC) [41]. A smaller DIC indicates a better fit and a difference between models of 10 or more is a strong indicator of a better model.

The Bayesian model was fitted in JAGS version 4.3.0 [37]. We used two chains each with 5,000 samples thinned by 5 with a burn-in of 5,000. We visually checked the convergence and mixing of the two chains. The odds ratios with 95% credible intervals for the three models are presented in Table 3. The Bayesian p-values for Model 3 are presented in the last column of Table 3.

#### Heterogeneity

Figure 3 in the main text presents the interaction effects for Gender, Race and Education. Equation 1 presents the interaction model that was specified for Gender, Equation 2 for Race, Equation 3 for Education, and Equation 4 for state % Trump votes. The estimated odds ratios with 95% credible intervals for the Gender, Race and Education interaction models are presented, respectively, in Table 4, Table 5, Table 6, and Table 7.

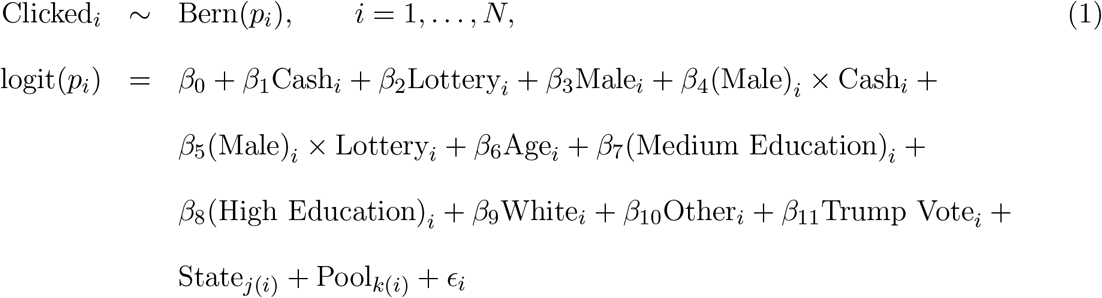

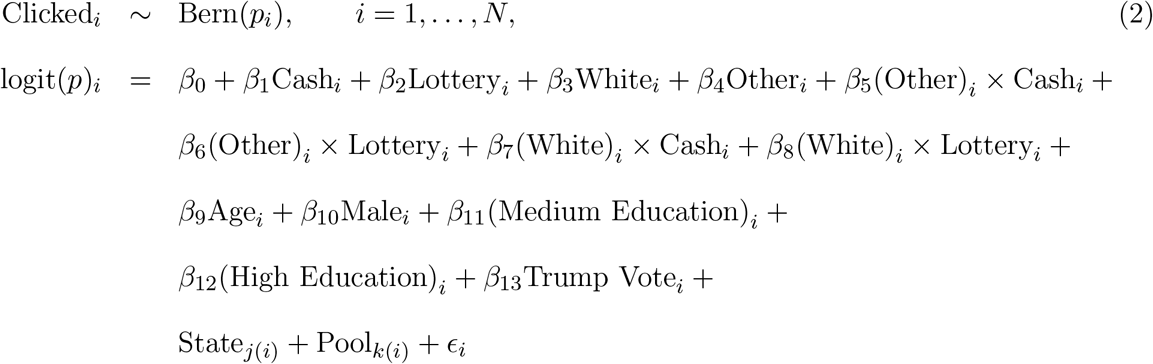

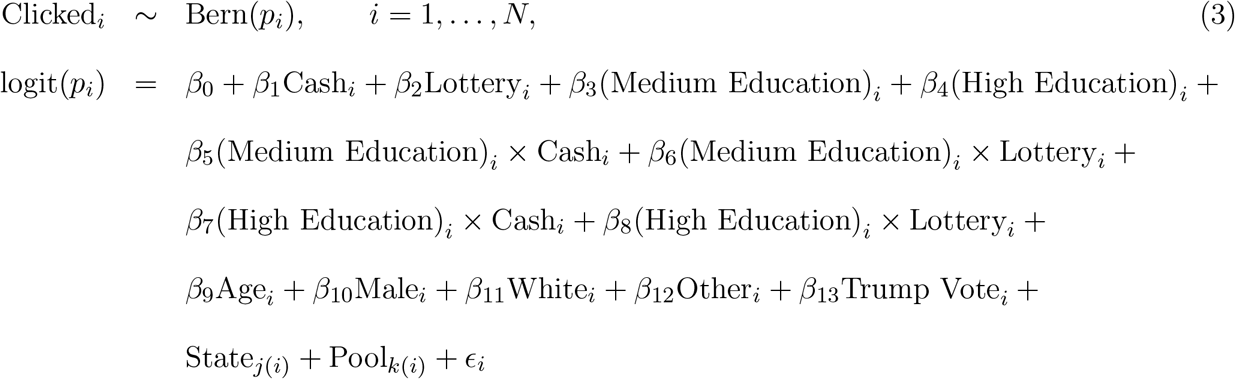

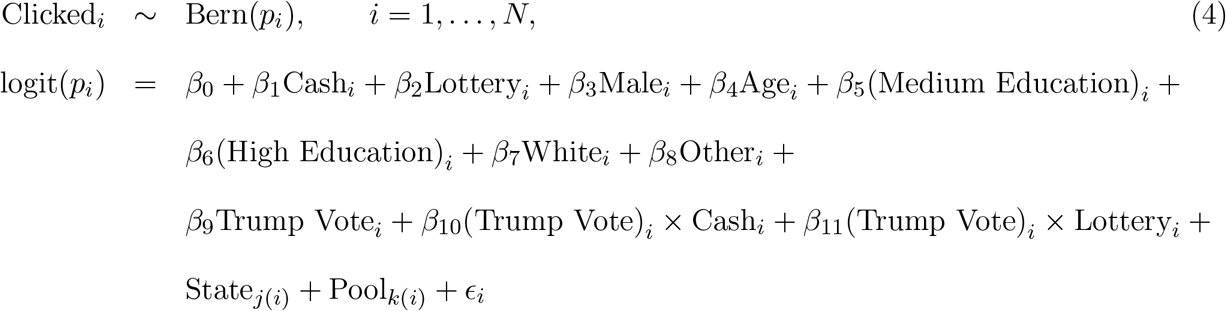

**Table 7:** Odds ratios and 95% credible intervals for the multiple logistic regression model examining an interaction between state Trump vote and treatment

| Variable | OR | CI | p-value |
| --- | --- | --- | --- |
| Cash Voucher Incentive | 1.53 | 1.10 to 2.14 | 0.012 |
| Lottery Incentive | 0.82 | 0.58 to 1.16 | 0.267 |
| Gender = Male | 0.93 | 0.69 to 1.24 | 0.630 |
| Trump × cash voucher | 1.14 | 0.77 to 1.69 | 0.508 |
| Trump × lottery | 0.86 | 0.57 to 1.31 | 0.488 |
| Age (+10 years) | 1.19 | 1.07 to 1.32 | <0.001 |
| Race = Other | 0.80 | 0.44 to 1.42 | 0.454 |
| Race = White | 0.68 | 0.47 to 0.98 | 0.040 |
| Education = Low | 1.33 | 0.88 to 2.02 | 0.176 |
| Education = Medium | 0.97 | 0.65 to 1.47 | 0.873 |
| State % Trump vote (+10%) | 1.15 | 0.86 to 1.57 | 0.360 |

All four models include the random effects of state and pool defined using Normal distributions with vague gamma priors for the inverse-variance:

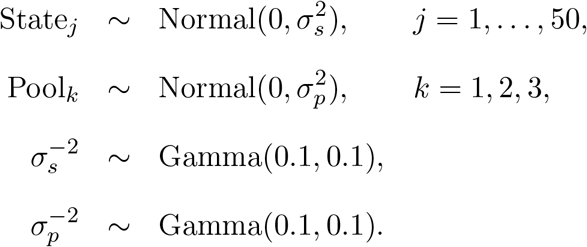

## Data Availability

Replications files and data from this project will be made available at the time of publication in a peer-review journal from https://dataverse.harvard.edu/dataverse/VaccineIncentives

## Footnotes

1 The OSF Registries pre-registration, U.S. COVID-19 Vaccine Incentives Experiment: 10.17605/OSF.IO/ADRW3

2 In our pre-registration calculations we established a sample size objective of 500 respondents in each of the three treatment groups. Due to the adaptive nature of the random assignment algorithm we recruited a total of 8% more subjects than anticipated.

3 We build on the approach, and the R code, made available by Moore and Moore [29], Moore [28], Moore and Schnakenberg [27], Cavaille [7]

4 Treatment 1 (control) video: https://youtu.be/V3DUCC8xnD0. Treatment 2 video: https://youtu.be/QIm_vbpe_go. Treatment 3 video: https://youtu.be/gF045EaPj-o.

5 The screen shots of this text are available in the Online Supplemental Material.

6 Diag Davenport describes how to deploy the API and embed it into a Qualtric’s survey https://diagdavenport.com

